# Medicaid Home and Community Based Services are vital for adults with intellectual and developmental disabilities

**DOI:** 10.1101/2025.09.10.25335499

**Authors:** Samuel B Rosenberg, Josue Antonio Garcia Estrada, Salina Tewolde, Alianna Higgins, Junjun Tao, A Alex Levine, Emily K Q Sisson, Amy Michals, Eric Rubenstein

## Abstract

Home and Community Based Services (HCBS) are Medicaid funded services that support independence, person-centered care, and connection to community for disabled people enrolled in Medicaid. With drastic Medicaid cuts on the horizon due to the 2025 Budget Reconciliation Bill, HCBS will likely be impacted. Our objective was to describe HCBS use by service type category among all adults enrolled in Medicaid with intellectual and developmental disabilities (IDD) diagnoses in 2022. We evaluated differences by state, IDD type, and race and ethnicity. We used Medicaid data from all adult enrollees with claims for autism, intellectual disability and Down syndrome and identified HCBS use using established algorithms. Of 1,519,852 adults with IDD, 63.1% (N=958,437) received any HCBS in 2022. The most used HCBS types were case management (32.5%), home based services (32.0%) and non-medical transportation (18.2%). There were limited differences by race and ethnicity, even after adjustment by state. With large Medicaid cuts forthcoming HCBS are optional services that are not mandated to be covered and a possible area for cuts that will disproportionately harm the IDD community.

## Introduction

Home and Community Based Services (HCBS) are Medicaid funded services that support independence, person-centered care, and connection to community for disabled people enrolled in Medicaid.^1^ In contrast to usual Medicaid covered services, these services are delivered in the home or community with the goal of addressing needs related to activities of daily living. Because HCBS are not a mandatory Medicaid benefit, each state determines which services it will cover, who is eligible, how someone is enrolled, and how the program is administered, often leading to within and between state variability in who receives services.^2, 3^ There has been extensive study of outcomes of HCBS, with HCBS receipt being associated with reduced days of inpatient hospitalizations and emergency department visits, more community living, and more preventative care.^4^

Adults with IDD enrolled in Medicaid are a population that are highly reliant on HCBS to maintain living in their community. IDD are conditions that present at birth and are characterized by significant limitations in intellectual functioning and adaptive behavior.^5, 6^In previous generations, people with IDD were excluded from their communities and placed in institution.^7^ With the Supreme Court’s *Olmstead vs. L*.*C*. ruling in 1999, states must provide the most integrated setting to meet a disabled person’s needs^8^; HCBS are one way that states do that. Now, over 1,000,000 people with IDD are enrolled in Medicaid HCBS waivers,^9^ the predominant pathway for states’ to provide tailored HCBS (e.g., personal care assistance, case management, transportation, and support self-directing care) to populations such as older adults and people with IDD.

Because of the state variability in HCBS programs,^9^ it is difficult to characterize HCBS use in the U.S. population. There have been recent advances in Medicaid data quality through the Transformed Medicaid Statistical Information System (T-MSIS) which has enabled exploration of HCBS use.^10, 11^ CMS has developed an algorithm to use claims, service locations, type of service code, and state to document which type of HCBS an enrollee receives.^1, 11, 12^ They have created thirteen categories: home-based services, case management services, nursing services, round-the-clock services, supported employment services, day services, home delivered meal services, caregiver support services, services directing supporting participant directed services, participant training, non-medical transportation, community transition services, and technical modification or equipment. With this advance in HCBS data, we are beginning to see better understanding of how HCBS are used nationally.^13, 14^

With drastic Medicaid cuts on the horizon due to the 2025 Budget Reconciliation Bill,^15, 16^ HCBS will likely be impacted. While some home health services are required to be provided by states, the overwhelming majority of HCBS are optional.^15^ As funding concerns grow, states may turn to cutting these services. Yet, HCBS use has not been extensively described in the IDD population, preventing us from anticipating some of the effects of potential cuts. Therefore, our objective was to describe HCBS use by service type category among all adults enrolled in Medicaid with identified IDD in 2022. We evaluated differences by state, IDD type, and race and ethnicity.

## Methods

### Sample

We used 2022 T-MSIS Analytic Files data from the Down Syndrome Toward Optimal Trajectories and Health Equity using Medicaid Analytic eXtract (DS-TO-THE-MAX) cohort. The DS-TO-THE-MAX cohort includes beneficiaries ≥18 years with diagnoses of Down syndrome, autism, or intellectual disability enrolled in Medicaid at any point from 2011-2022, enrolled for at least 12 months. The [institution omitted] Institutional Review Board determined that the study was not human subject research and required no further review.

### IDD identification

We used a narrow definition of IDD (e.g. not including cerebral palsy on claims) as used by the American Association for Intellectual and Developmental Disability^5^ with well specified ICD-9 and ICD-10 codes. We used existing and validated algorithms to identify IDD, which required a diagnosis code on one inpatient claim or two or more other claim types in any year of enrollment.^17, 18^ We created non-exclusive binary variables for autism, Down syndrome, and intellectual disability diagnosis and an exclusive categorical variable (Down syndrome, autism without Down syndrome, intellectual disability without autism or Down syndrome). We further specified autism with intellectual disability and autism without intellectual disability to attempt to explore the diversity within the population of Medicaid enrollees with autism.

### HCBS identification

We identified HCBS categories using the methodology outlined in the TAF Methodology Brief and Supplemental 1.^*11*^ Categories were from the Medicaid Analytic eXtract HCBS taxonomy. We used Healthcare Common Procedure Coding System (HCPCS), ICD-10 Procedure Coding System (ICD-10-PCS), or Current Procedural Terminology (CPT) codes in the Other Services files and a multi-stage hierarchical approach to create exclusive categories. We prioritized state-specific procedure codes over generic, national codes because HCBS are implemented at the state level. For claims without state codes, we used national procedure codes to identify HCBS claims: either (1) using the procedure code only, (2) using the procedure code and other codes (e.g., type of service or procedure code modifiers), or (3) using the procedure code and codes indicating a service was paid by an HCBS program (program type, benefit type, HCBS code, or waiver type).

We used the thirteen HCBS categories created by CMS in the TAF Methodology brief. We created variables for any HCBS use in the year, any use of HCBS by category in the year, the number of months using any HCBS, and the number of months using a specific HCBS category. We calculated percentage of the year with HCBS by dividing HCBS months by total months enrolled. We could not tie HCBS type of HCBS program (e.g. 1915c) because individuals could have multiple HCBS program types which we could not differentiate which covered the service.

### Other covariates

Covariates included age on January 1, 2022, sex, race and ethnicity, region, and enrollment. We defined age in years both continuously and categorically. Categories for age group include 18-24, 25-34, 35-44, 45-54, 55-64 and 65 and older. Race was self-reported to state Medicaid agency and categorized by CMS into five categories Asian or Pacific Islander, Black, Multiple races, Native American, or White. Similarly, ethnicity was defined by CMS as non-Hispanic or Hispanic. To handle missing race and ethnicity data, we first examined up to 11 years of enrollment for an individual to examine race or ethnicity reporting in prior years. If they had ever reported race or ethnicity, we considered that their race or ethnicity in 2022. If they were still missing (∼12%), we used multiple imputation to probabilistically determine race (see Levine et al ^14^ for further description). We included variables for state (including Washington, D.C., Puerto Rico, and the U.S. Virgin Islands) and region (Northeast, South, Midwest, West, and U.S. Territories) lived in for the majority of 2022. Enrollment was defined monthly and over the year, as Medicaid-only or Dual enrollment in both Medicare and Medicaid.

### Statistical analysis

We estimated any HCBS use across all participants in the sample and any HCBS use by month. Denominators were the population with IDD enrolled in Medicare and/or Medicaid in 2022. We estimated any HCBS use and specific HCBS use by sex, race (most prevalent races white, Black, and Asian), ethnicity, and IDD type (autism, intellectual disability, Down syndrome; not exclusive). We evaluated any and specific HCBS use by state, accounting for the size of IDD population in the state.

To further understand differences by IDD type and race/ethnicity, we ran general linearized models clustered by state to calculate adjusted differences (in percentage points) in service use. In the model we adjusted for age (in categories) and dual enrollment. We ran IDD type and race/ethnicity in the same models since they should be adjusted for one another when estimating effects of either variable. For IDD type, we categorized as Down syndrome, autism without Down syndrome, intellectual disability without Down syndrome and without autism.

We calculated 95% confidence intervals for all estimates. Because of the large sample size, we did not use null-hypothesis testing as P values would be likely significant when not clinically meaningful. We use Stata 18, SAS 9.4, and R 4.4.2 for data analysis and visualization.

## Results

In 2022, there were 1,519,852 adults ≥18 years with IDD enrolled in Medicaid, 60.3% were male (N=916,023) and 39.7% were female (N=603,811) (Table 1). Mean age was 40.1 years (SD:16.6) and median age was 36.0 (Interquartile range: 26, 53) with 10.3% being older than 65 years (N=160,075). 69.8% were White, 20.2% Black, 3.4% Asian, 1.5% Pacific Islander, 0.9% Native American, and 4.3% had multiple races. By ethnicity, 83.4% were non-Hispanic White, and 16.6% were Hispanic. Among enrollees, 95.8% were enrolled for all 12 months of 2022 and 53.4% (N=816,695) were dual enrolled in Medicare for at least one month. By IDD type (not exclusive), 6.8% (N=103,673) had claims for Down syndrome, 37.4% (N= 569,151) had claims for autism, of which 45% [N=260,331] also had a claim for intellectual disability. Three-quarters (77.7% [N=1,180,740]) had claims for intellectual disability.

**Table 1.** HCBS Demographics 2022.

| HCBS Demographics 2022 |  |  |  |
| --- | --- | --- | --- |
| Overall | N | Yes HCBS (%) | No HCBS (%) |
|  | 1,519,852 | 958,437 (63.1) | 561,415 (36.9) |
| **Age Group** |  |  |  |
| 18-24 | 310,394 | 189,523 (61.1) | 120,871 (39.0) |
| 25-34 | 406,701 | 255,416 (62.8) | 151,285 (37.2) |
| 35-44 | 261,069 | 169,142 (64.8) | 91,927 (35.2) |
| 45-54 | 187,900 | 121,703 (64.8) | 66,197 (35.2) |
| 55-64 | 193,713 | 125,422 (64.7) | 68,291 (35.2) |
| 65 and older | 160,075 | 97,231 (60.7) | 62,844 (39.3) |
| **Cohort** |  |  |  |
| Autism | 569,151 | 344,679 (60.6) | 224,472 (39.4) |
| Intellectual Disability | 260,331 | 195,216 (75.0) | 65,115 (25) |
| Down syndrome | 103,673 | 73,560 (71.0) | 30,113 (29) |
|  | 1,180,740 | 792,987 (67.2) | 387,753 (32.8) |
| **Race** |  |  |  |
| White | 1,028,181 | 647,427 (63.0) | 380,754 (37.0) |
| Black | 296,986 | 181,769 (61.2) | 115,217 (38.8) |
| Pacific Island | 22,493 | 13,208 (58.7) | 9,285 (41.3) |
| Asian | 49,607 | 34,367 (69.3) | 15,240 (30.7) |
| Native American | 12,774 | 8,380 (65.6) | 4,394 (34.4) |
| Multiple Races | 63,410 | 42,923 (67.7) | 20,487 (32.3) |
| **Ethnicity** |  |  |  |
| Non-Hispanic | 1,229,246 | 775,843 (63.1) | 453,403 (36.9) |
| Hispanic | 244,205 | 152,231 (62.3) | 91,974 (37.7) |
| **Region** |  |  |  |
| Northeast | 355,415 | 206,051 (58.0) | 149,364 (42.0) |
| South | 369,528 | 230,966 (62.5) | 138,562 (37.5) |
| Midwest | 472,291 | 283,103 (59.9) | 189,188 (40.1) |
| West | 315,581 | 237,189 (75.2) | 78,392 (24.8) |
| US Territories | 7,037 | 1,128 (16.0) | 5,909 (84.0) |
| **Enrollment** |  |  |  |
| Medicaid | 1,518,850 | 958,178 (63.1) | 560,672 (36.9) |
| Dual | 816,695 | 509,975 (62.4) | 306,720 (37.6) |

In our population, 63.1% of adults with IDD (N=958,437) used any HCBS in 2022. HCBS use was consistent across age groups, ranging from 60.7% among those 65 and older to 64.8% among 35-44 and 45-54 years. By IDD-type, enrollees with Down syndrome (71.0%) and intellectual disability (67.1%) were more likely to use any HCBS compared to autistic enrollees (60.6%). Among enrollees who used HCBS, 69.8% were White, 19.6% Black, 1.4% Pacific islander, 3.7% Asian, 0.9% Native American, and 4.6% had multiple races. By ethnicity, 83.6% were non-Hispanic White, and 16.4% were Hispanic. I. Among HCBS users, 517,793 (54.0%) used HCBS in all months they were enrolled in Medicaid in 2022 (Supplemental 2).

Across U.S. states and territories, rates of HCBS use in 2022 ranged from 29.5% (95% CI: 28.5%, 30.5%) to 91.6% (95% CI: 91.0%, 92.3%) (Figure 1). Except for six states, more than 50% of the IDD population across these areas have received at least one HCBS in the year. The highest proportion of Medicaid enrollees with IDD who received HCBS were in the West and South. Receipt of any HCBS across racial and ethnic groups by state did not meaningfully differ (Supplemental 3). For each state, individuals with an autism diagnosis only had significantly lower HCBS use rates while those with Down syndrome or those with co-occurring autism and intellectual disability had the highest rates of HCBS use (Supplemental 4).

**Figure 1.**
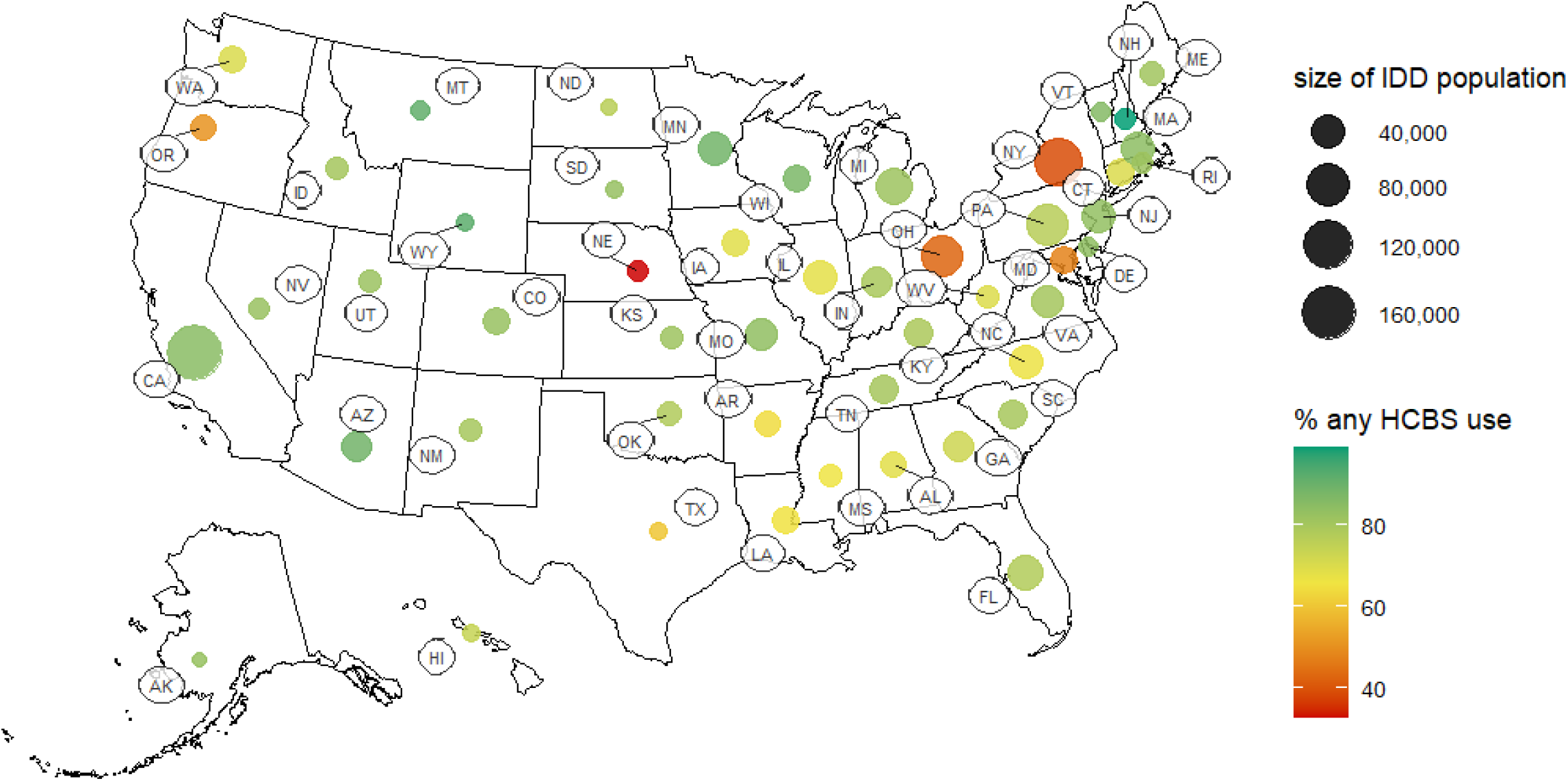
Any HCBS use by state and count of adults with intellectual and developmental disability enrolled in Medicaid, 2022 IDD: intellectual and developmental disability HCBS: Home and Community Based Service

The most used HCBS types were case management (32.5%), home-based services (32.0%) and non-medical transportation (18.2%; Figure 2). Less than 1/10^th^ of a percent used community transition services and 1.0% received home delivered meals. Only 28.6% of people with claims for autism had home-based services use compared to 34.9% for the intellectual disability group and 38.8% for the Down syndrome group. Those with Down syndrome claims had the highest rate of day services and caregiver support service use. By race, Asian individuals were most likely to receive home-based care (42.8%) compared to white (30.6%) and Black (33.0%) peers and had a 10%-point higher use of caregiver support compared to the white and Black groups (15% compared to 5%). By ethnicity, the largest difference was in case management, with 34.6% of non-Hispanic enrollees with IDD receiving the service compared to 22.0% of Hispanic enrollees with IDD. Service type used by state is presented in Table 2, illustrating the heterogeneity across the country.

**Table 2.** Ever use of specific Home and Community Based Service types, overall and by demographic characteristics, for adults with intellectual and developmental disabilities enrolled in Medicaid in 2022.

|  | **Autism** | **Intellectual disability** | **Down syndrome** | **White** | **Black** | **Asian** | **Hispanic** |
| --- | --- | --- | --- | --- | --- | --- | --- |
|  | N=559,291 | N=1,143,316 | N=101,080 | N=1,028,181 | N=296,986 | N=49,607 | N=244,205 |
| **Any HCBS** | 334,295 (60.5%) | 766,588 (67.1%) | 71,740 (71.0%) | 647,427 (62.3%) | 181,769(61.2%) | 34,367 (69.3%) | 152,131 (62.3%) |
| **Service type** |  |  |  |  |  |  |  |
| Home-based services | 157,901 (28.6%) | 399,035 (34.9%) | 39,261 (38.8%) | 314,375 (30.6%) | 98,015 (33.0%) | 21,254 (42.8%) | 86,253 (35.3%) |
| Case management | 191,296 (34.6%) | 394,956 (34.5%) | 35,370 (35.0%) | 357,714 (34.8%) | 91,511 (30.8%) | 10,924 (22.0%) | 53,768 (22.0%) |
| Nursing | 24,458 (4.4%) | 69,447 (6.1%) | 5,483 (5.4%) | 52,279 (5.1%) | 17,507 (5.9%) | 2,236 (4.5%) | 12,935 (5.3%) |
| Round the clock | 69,568 (12.6%) | 180,384 (15.8%) | 14,820 (14.7%) | 152,481 (14.8%) | 29,392 (9.9%) | 3,931 (7.9%) | 25,214 (10.3%) |
| Supported employment | 18,892 (3.4%) | 53,522 (4.7%) | 4,222 (4.2%) | 45,575 (4.4%) | 9,955 (3.4%) | 1,498 (3.0%) | 8,106 (3.3%) |
| Day services | 78,411 (14.2%) | 209,065 (18.3%) | 22,900 (22.7%) | 169,225 (16.5%) | 38,132 (12.8%) | 8,928 (18.0%) | 35,413 (14.5%) |
| Home delivered meals | 3,821 (0.7%) | 12,719 (1.1%) | 853 (0.8%) | 9,255 (0.9%) | 4,460 (3.4%) | 238 (0.5%) | 606 (0.2%) |
| Caregiver support | 41,022 (7.4%) | 75,642 (6.6%) | 11,123 (11.0%) | 57,193 (5.6%) | 17,354 (5.8%) | 7,690 (15.5%) | 25,149 (10.3%) |
| Participant directed | 23,064 (4.2%) | 43,698 (3.8%) | 5,893 (5.8%) | 42,232 (4.1%) | 7,706 (2.6%) | 1,363 (2.7%) | 5,306 (2.2%) |
| Participant training | 35,251 (6.4%) | 73,364 (6.4%) | 5,728 (5.7%) | 66,596 (6.5%) | 14,947 (5.0%) | 3,255 (6.6%) | 4,842 (2.0%) |
| Non-medical transportation | 84,433 (15.3%) | 233,192 (20.4%) | 19,818 (19.6%) | 187,592 (18.2%) | 51,361 (17.3%) | 9,702 (19.6%) | 14,358 (5.9%) |
| Community transition | 464 (0.1%) | 964 (0.1%) | 28 (0.03%) | 870 (0.08%) | 227 (0.08%) | 23 (0.05%) | 133 (0.05%) |
| Technical modifications | 71,302 (12.9%) | 214,242 (18.7%) | 19,978 (19.8%) | 179,802 (17.5%) | 46,706 (15.7%) | 6,527 (13.2%) | 37,730 (15.5%) |

**Figure 2.**
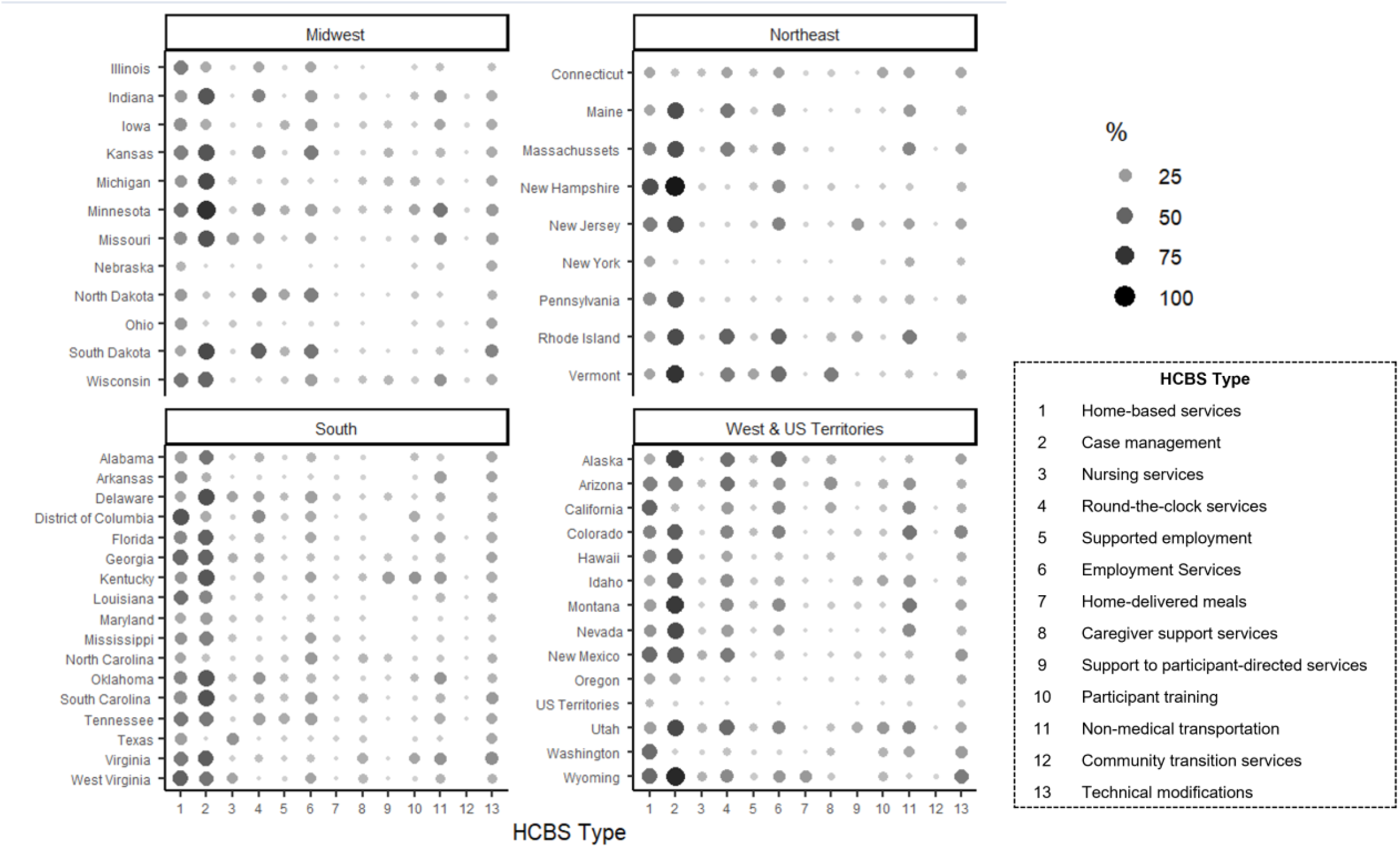
Home and Community Based Service use by type and state among Medicaid enrollees with intellectual and developmental disabilities, 2022

After adjusting for potential confounders (Table 3), individuals with Down syndrome were 6.9 percentage points (95% CI: 3.4, 10.4) more likely to use HCBS compared to those in the intellectual disability group. Those with autism had utilization rates 3.8 percentage points (95% CI: −7.5, 0.0) less than the intellectual disability group. the biggest difference in services used were people with Down syndrome being more likely to receive home-based services and day services compared to those with intellectual disability or autism. We did not find significant differences in overall HCBS use by race and ethnicity (95% confidence interval contained the null). Asian enrollees with IDD were 12.3 percentage points (95% CI: 2.5, 22.2) more likely to receive caregiver support and home-based services (7.3 percentage points, 95% CI: 0.5, 14.2) compared to white non-Hispanic enrollees. Hispanic enrollees were 14.3 percentage points less likely to receive case management compared to white enrollees (95% CI: −23.8, −4.9).

**Table 3.** Adjusted differences in percentage points of Home and Community Based Services used among people with intellectual and developmental disabilities enrolled in Medicaid in 2022, by disability type and race/ethnicity Home delivered meals and community transition services were excluded due to low sample size.

|  | **Intellectual**<br>**disability**<br>**N=1,143,316** | **Autism**<br>**N=559,291**<br>**% D (95% CI)** | **Down**<br>**syndrome**<br>**N=101,080**<br>**% D (95% CI)** | **White**<br>**N=1,028,181** | **Black**<br>**N=296,986**<br>**% D (95% CI)** | **Asian**<br>**N=49,607**<br>**% D (95% CI)** | **Hispanic**<br>**N=244,205**<br>**% D (95% CI)** |
| --- | --- | --- | --- | --- | --- | --- | --- |
| **Any HCBS** | REF | -3.8 (-7.5, 0) | 6.9 (3.4, 10.4) | REF | -1.9 (-4.1, 0) | 5.9 (-2.5, 14.4) | -1.5 (-9.2, 6.2) |
| **Service type** |  |  |  |  |  |  |  |
| Home-based services | REF | -3.6 (-5.8, -1.4) | 6.3 (3.4, 9.3) | REF | 2.2 (0, 4.5) | 12.3 (2.5, 22.2) | 4.9 (-2.8, 12.7) |
| Case management | REF | 1.8 (-2.6, 6.2) | 3.5 (-0.5, 7.4) | REF | -3.7 (-8.1, 0.7) | -10.8 (-22.2, 0.6) | -14.3 (-23.8, -4.9) |
| Nursing | REF | -1.5 (-2.0, 0.0) | -0.5 (-1.6, 0.7) | REF | 0.8 (-0.4, 2.0) | -0.6 (-0.3, 0.1) | 0.0 (-3.0, 3.2) |
| Round the clock | REF | 0.6 (-1.1, 2.4) | 1.2 (-1.3, 3.8) | REF | -4.8 (-7.6, -1.9) | 2.0 (-5.6, 2.1) | -4.1 (-0.8, 0.0) |
| Supported employment | REF | -0.8 (-1.9, 0.3) | -1.2 (-2.0, 0.0) | REF | -1.0 (-2.0, 0.0) | -0.9 (-1.9, 0.1) | -1.1 (-0.3, 0.6) |
| Day services | REF | -0.4 (-3.4, 2.7) | 6.6 (3.7, 9.6) | REF | -3.0 (-5.2, -0.8) | 2.4 (-3.8, 8.6) | -1.7 (-6.7, 3.3) |
| Caregiver support | REF | -0.9 (-2.4, 0.5) | 4.2 (2.5, 5.9) | REF | 0.3 (-0.8, 1.4) | 7.3 (0.5, 14.2) | 4.0 (-2.1, 10.0) |
| Participant directed | REF | 0.0 (-1.0, 1.1) | 2.4 (0.7, 0.4) | REF | -1.3 (-2.3, -0.1) | -0.8 (-3.5, 1.8) | -1.3 (-4.3, 1.6) |
| Participant training | REF | 0.2 (-0.5, 0.8) | -0.5 (-1.6, 0.7) | REF | -1.2 (-2.4, 0.0) | 0.1 (-1.1, 1.4) | -0.3 (-3.5, 2.8) |
| Non-medical transportation | REF | -3.6 (-5.8, -1.4) | -0.4 (-3.0, 0.2) | REF | -1.4 (-3.9, 1.1) | 1.4 (-3.7, 6.4) | 0.4 (-4.5, 5.2) |
| Technical modifications | REF | -4.9 (-5.6, -4.1) | 1.7 (0.5, 2.9) | REF | -2.3 (-3.5, -1.2) | -4.4 (-6.3, -2.6) | -2.2 (-3.7, -1.2) |

Models additionally adjusted for age and dual enrollment in Medicare at the individual level and clustered by state.
Intellectual disability, autism, and Down syndrome were categorized as exclusive, with Down syndrome and co-occurring autism being in the Down syndrome group and autism with co-occurring intellectual disability being in the autism group.
CI: Confidence interval

## Discussion

Disabled people have care needs that exist beyond the scope of typical healthcare system. HCBS are designed to address those needs to keep disabled people in their communities and prevent or delay transition to long-term care. Based on 2022 Medicaid claims, we found that around 60% of enrollees with IDD used any HCBS, most prevalent being case management and home-based services. Our findings have major implications for the IDD population in a time of impending Medicaid funding cuts.

We found more than half of adults with IDD identified in Medicaid claims received any HCBS in 2022. This is aligned with our previous work documenting high rates of 1915(c) waiver receipt in 2016-2019^14^; however, the present study included services provided under different waiver types and programs. Depending on the state, Medicaid enrollees may be automatically enrolled in HCBS programs, or they may have to navigate burdensome eligibility and application processes.^19, 20^ Further, 2022 was during the Covid-19 Public Health Emergency which enabled states to strengthen and expand access to HCBS.^21^ Many states maintained these changes post public health emergency, but it could be expected that the amount of these services decreased and continue to decrease since 2023.^21^

We found few demographic differences in overall utilization and by service types. The differences we did see may be attributable to residual state differences and states’ HCBS policies. While the lack of major differences is promising, it does not tell us about differences in quality of care received or remaining unmet needs, which may be different by race, ethnicity, or age.^22^ There were some differences when comparing by intellectual and developmental disability type, which makes sense given the different functional limitations between conditions. Further, not all with IDD will receive Medicaid through disability pathways, which are necessary to receive HCBS. People with autism are more likely than those with intellectual disability or Down syndrome to receive Medicaid via income rather than disability^23-25^ and thus may have lower rates of HCBS use.^23-25^

Our study was limited by potential reporting issues in claims data. The algorithm we used to identify HCBS relies on procedure codes being reported accurately and if they are not accurately reported they may not be classified into the correct service category. Our objective was to describe service; therefore, state level data need to be interpreted in the context of state policies, demographics, and HCBS funding mechanisms. We used a narrow definition of IDD which covers much of the broader IDD population with more precision in ICD codes. Including other IDD that do not co-occur with intellectual disability or autism would give us a more complete picture of HCBS use. We used race and ethnicity variables provided by CMS and more refined categories would have helped us uncover more patterns. Nevertheless, we were able to use a full Medicaid sample of enrolled adults with IDD and identify HCBS. We used novel HCBS categories to understand patterns in service use and we identified differences by state, IDD type, and race ethnicity.

## Conclusion

There are major policy implications given our evidence of wide HCBS use. With future spending cuts to Medicaid passed in 2025,^16^ HCBS are at risk. Cuts will change how Medicaid is financed which may put states at risk and force them to decrease spending. If spending needs to decrease HCBS are optional services that are not mandated to be covered and a possible area for cuts.^22^ Additionally, there is already a shortage of personal care attendants who work long hours for little pay. These workers are often reliant on Medicaid via Medicaid expansion and are at risk for losing their health insurance if they remain in these types of jobs.^26^ Loss of these workers will further limit availability of needed HCBS. The data show that most people with IDD regularly use HCBS to maintain their lives in their community and future Medicaid cuts will likely disrupt these services.

## Data Availability

The data used in this study are Medicaid claims data obtained from the Centers for Medicare & Medicaid Services (CMS) under a data use agreement. These data are not publicly available due to privacy and legal restrictions. Access to the data may be granted to qualified researchers through application to CMS via the Research Data Assistance Center (ResDAC) and completion of the necessary data use agreements.

https://resdac.org/

