## Supplemental 2-4 for "Medicaid Home and Community Based Services are vital for adults with intellectual and developmental disabilities"

Supplement 2. Percentage of months enrolled that Home and Community Based Services were used among Medicaid enrollees with intellectual and developmental disabilities, 2022

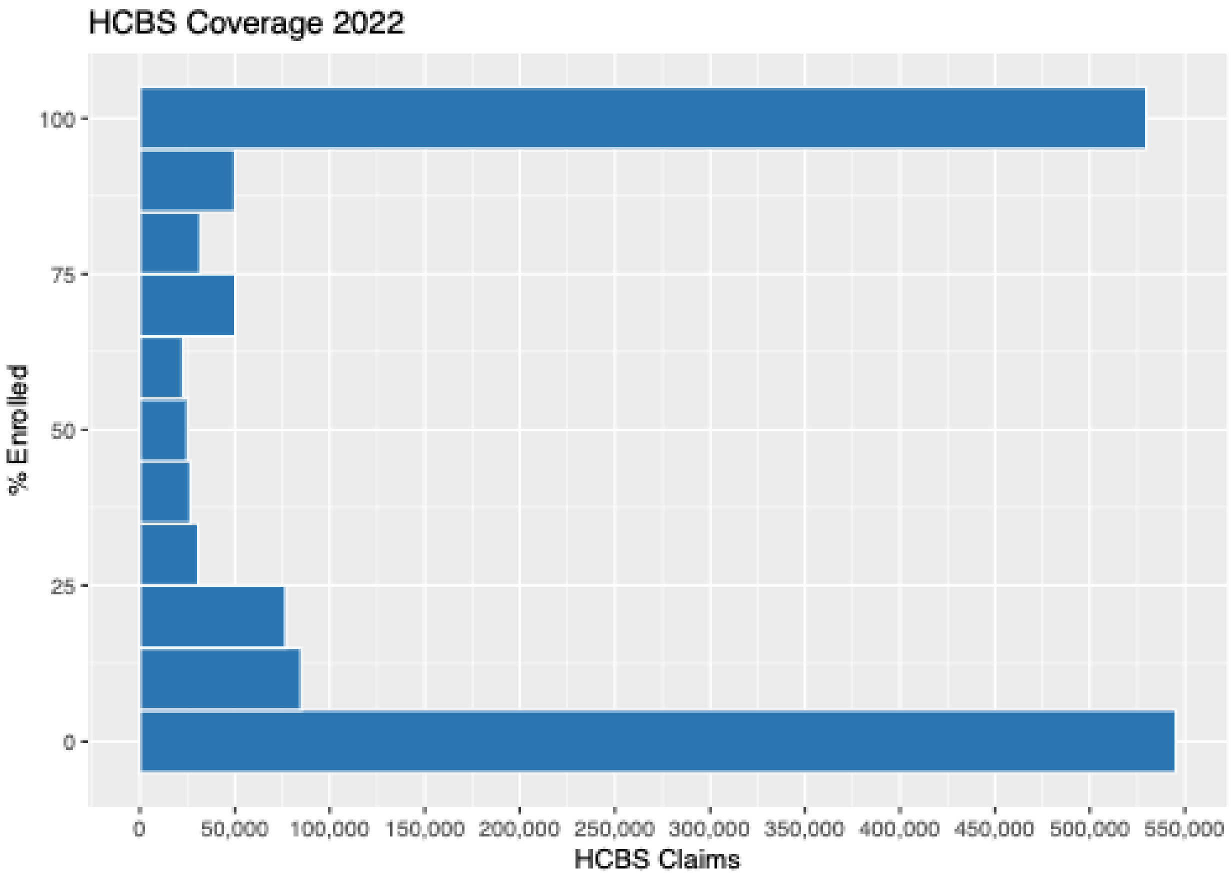

Supplement 3. Home and Community Based Service use by race and state among adults with intellectual and developmental disabilities, 2022

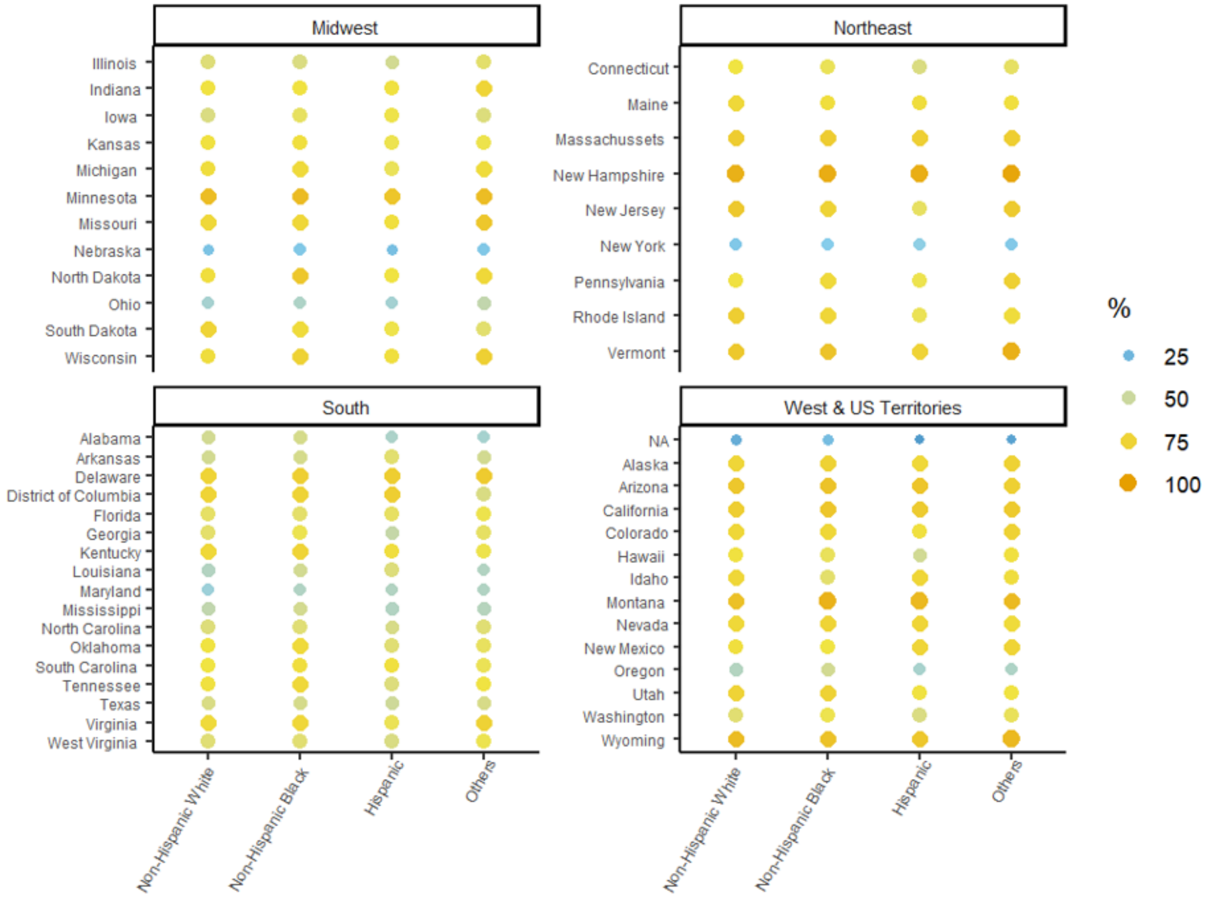

Supplement 4. Home and Community Based Service use by intellectual and developmental disability type among Medicaid enrollees with intellectual and developmental disabilitis, 2022

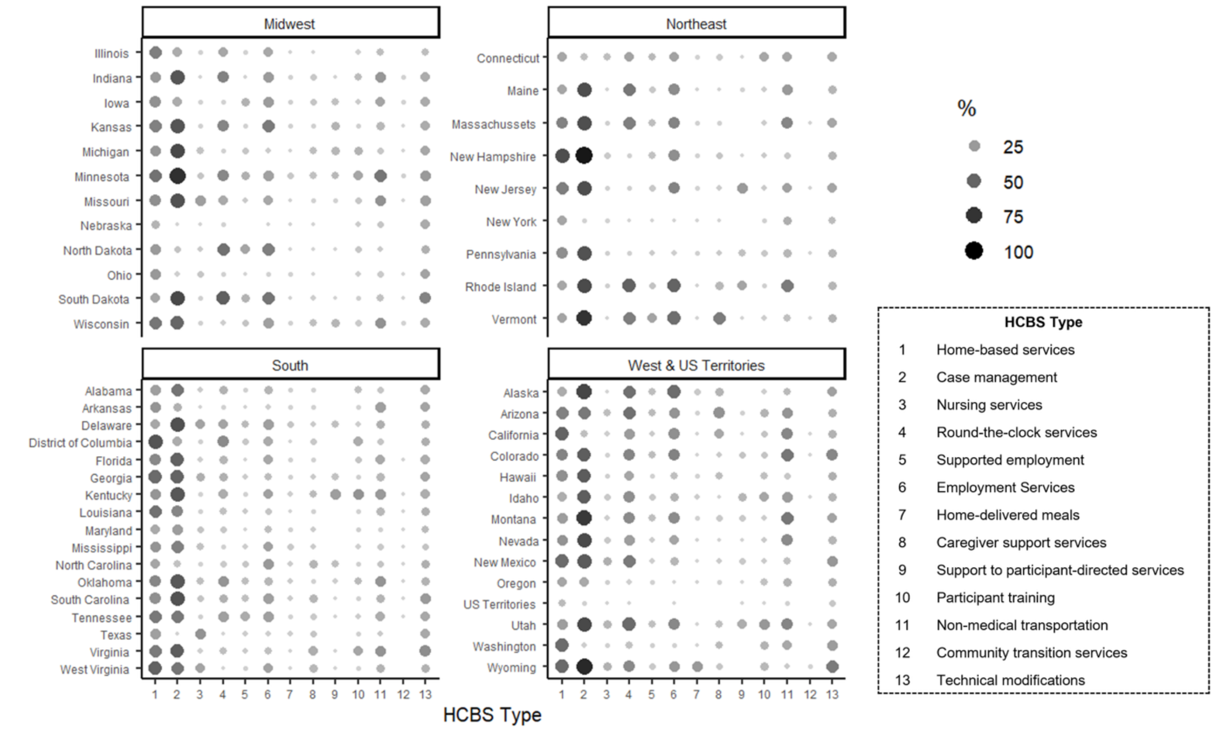
